# Spatial Association of Respiratory Health with Social and Environmental Factors: Case Study of Cook County, IL

**DOI:** 10.1101/2021.04.29.21256319

**Authors:** Aynaz Lotfata, Alexander Hohl

**Affiliations:** Department of Geography, Chicago State University, Chicago, IL, 60628, USA; Department of Geography, The University of Utah, Salt Lake City, UT, 84112, USA

**Keywords:** Social and Environmental Factors, Respiratory Health, Asthma, Spatial Analysis

## Abstract

**Background:** People who live with respiratory diseases like asthma are more likely at risk of serious illness. Spatial analytic techniques allow for discovering areas of concern and finding correlates of asthma prevalence.

**Objective:** There is growing interest in disentangling the impacts of socioeconomic and environmental factors on respiratory health, their spatial correlation, and the demographic profile of people at risk of respiratory diseases. It is important to know how people with asthma are geographically distributed and what social and environmental factors correlate with asthma. Thereby, the purpose of the study is to describe socioeconomic factors associated with asthma prevalence in Cook County, IL and to identify the significant risks and the protective factors to control asthma.

**Data and Methods:** Data obtained from CDC 2018 SVI, ACS, the City of Chicago Data Portal, HealthData.gov, and ESRI. In this paper, we illustrate the usefulness of geospatial regression analysis in the analysis and presentation of spatially distributed asthma prevalence among the population with disabilities, minorities with the language barrier, nonwhite population, age 17 and younger, and age 65 and older in the census tracts of Cook County, IL where Chicago Metropolitan Area located. In addition, we map the spatial variation of asthma prevalence with variation in the tree canopy, access to medical centers, air quality, and household quality. Lastly, we used bivariate mapping to illustrate the spatial distributions of residential land use and tree covers.

**Results:** Our findings show a good correlation between asthma and socioeconomic and physical factors including age 17 and younger, age 65 and older, population with disabilities, a minority with the language barrier, tree canopy, access to medical centers, air quality, and household quality. The aged 65 and older, 17 and younger, and people with disabilities are found to have a higher asthma prevalence in areas around the industrial corridors in southeast and west sides of Cook County, IL. Results may guide further decisions in planning for asthma research and intervention, especially for identifying vulnerable areas and people.

## Background

Since the early 1990s, respiratory disease burden has been recognized as a national public health concern in the United States (Department of Health and Human Services, 2000). The US Environmental Protection Agency (EPA) and American Public Health Association (APHA) reported urbanization as a contributor to respiratory diseases (Wong & Chow, 2008). Asthma, a non-communicable respiratory disease, is increasing worldwide. Current asthma rates among urban populations remain high despite improvements in available therapies and medicines (Van Dorn, 2017) and affect individuals of certain racial/ethnic backgrounds disproportionately. The World Health Organization (WHO) estimates 417,918 deaths due to asthma at the global level in 2016 (WHO, 2018). In recent years, asthma rates have risen so much in the US that medical and public health professionals invariably speak of it as a new epidemic. The number of individuals with asthma in the United States is more than 25 million (CDC, 2020) and approximately 850,000 people with asthma are in Illinois (IDPH, 2020). Therefore, the identification of risk factors responsible for this increase is a crucial public health practice.

There is a growing recognition that social determinants of health - the conditions in which people live, learn, work, play and worship - affects wellbeing and produces disparities (Thornton et al., 2016; Schroeder, 2007; Wilkinson & Marmot, 2003). Despite the medical advancement, inequity in healthcare access, inadequate treatment, poor housing, social and psychological stressors, lifestyle, and socioeconomic status are linked with an uneven burden in asthma mortality in urban populations.

Access to and use of asthma health services affect asthma management and control among older adults and people with disabilities (Rodriguez et al., 2019; Curto et al., 2019; Shah & Newcomb, 2018; Ponte et al., 2016; Davis, 2016; Jandasek et al., 2011). Health care should be affordable, enabling patients to access the treatments they need to live healthy (NHC, 2020). In neighborhoods with a high rate of crime, people have trouble accessing safe transportation (Mitchell & Murdock, 2005). Besides, effective communication between asthma patients and healthcare contributes to asthma diagnosis. It was found African American and Latino patients use different words to explain asthma symptoms (Bryant-Stephen, 2009; Hardie et al., 2000). Despite living in high pollution areas, asthma prevalence did not differ among the Hispanic Whites when compared with African-Americans and non-Hispanic Whites. Because the Hispanic population is bound to its ethnic groups and the Spanish language (Thakur et al., 2019). Poor English language limits access to high-quality care (even when they are insured).

Further, minority and low-income groups, with African American and Latino children who live in poor urban environments experiencing higher asthma morbidity and mortality than white children (Bryant-Stephens, 2009). Additionally, specific to asthma, family structure, including single-parent households were associated with asthma-related admission which is more than twice as likely admitted from married parents (Weinstein et al., 2019; Moncrief et al., 2014). Hur et al. (2015) found the relationship between single parent depression and asthma outcomes in children. Therefore, socioeconomic status, lifestyles, and access to medical care might impact asthma incidence and prevalence.

Several environmental disparities have been reported to be associated with asthma incidence, including impacts of exposure to air pollution (Anto, 2012; The Lancet Planetary Health, 2017), as manufacturing industrial corridors release more than 1 million pounds of toxic chemicals (e.g. cadmium, naphthalene and ethyl benzene) into the air every year (Illinois Annual Air Quality Report, 2017). Additional factors associated asthma are overcrowding multi-unit structures with poor indoor ventilation (Jie et al., 2011; Northridge et al., 2010), housing density, neglected and old buildings which are triggers for indoor allergies due to excessive moisture and pests, mobile homes (Qi Gan et al., 2017), and lack of green spaces (Alcock et al., 2017).

Studies have found a positive association between the amount of neighborhood tree cover and better overall health, primarily mediated by lower overweight/obesity and better social cohesion, and to a lesser extent by less type 2 diabetes, high blood pressure, and asthma. Alcock et al. (2017) showed that both trees and greenspace were, on average, associated with lower area rates of people with asthma hospitalizations. Trees and nature contribute to reducing air pollution improving respiratory health in urban areas (Soyiri & Alcock, 2018; Ulmer et al., 2016; Sullivan et al., 2014). Urban trees could buffer harmful exposures such as air pollution from traffic. Though the respiratory system has remarkable resilience to air pollution, constant exposure to elevated particle pollution will contribute to reduced respiratory function, even in healthy people (EPA, 2020). WHO reviews the health benefits of urban green spaces in four ways: improved air quality, increased physical activity, stress reduction, and greater social cohesion (Hrala, 2016). This mindset calls for interdisciplinary studies on the relation between the population’s respiratory health and spending time in nearby urban trees and green spaces. This also addresses a policy issued by the American Public Health Association (APHA, 2019) entitled, “Improving Health and Wellness through Access to Nature.”

Social and environmental interventions to reduce the disparity in respiratory health and asthma prevalence in urban areas are very complex. This gets important when asthma is a risk to public health. People with asthma are more likely at the risk of severe respiratory infections during the COVID-19 pandemic than are people without asthma (Liu et al., 2020; Dong et al., 2020; Corne et al., 2002).

Geospatial analysis can guide such interventions to prioritize locations for targeted interventions and resource allocation and identify vulnerable districts with elevated asthma prevalence. Spatial clustering techniques can supplement basic disease maps by identifying areas of statistically elevated asthma prevalence, therefore distinguishing whether the observed patterns occurred by chance or not (Kulldorff, 1997). Spatial regression is a popular technique to analyze geographic data that may violate the assumptions of ordinary least-squares (OLS). It accounts for spatial autocorrelation within the target variable and can reduce spatial trends in model residuals (Anselin, 2001). Hence, geospatial analysis is important for health officials and city planners to make informed decisions to reduce health disparities.

Cook County, IL includes the Chicago Metropolitan Area and exhibits an average 36 national rank in asthma prevalence (AAFA, 2020). In the last decade, rising hospitalization rates due to asthma have become a significant public health problem in Chicago (Thomas & Whitman, 1999). In recent years, more than half (58%) of all children with asthma in Chicago had a severe asthma attack (Respiratory Health Association of Metropolitan Chicago, 2018). Thereby, the purpose of this study is to examine the overall burden of asthma prevalence from the perspective of geospatial variation in recent years in Cook County.

### Purposes

- To describe geographical location of the population with asthma
- To identify areas of elevated asthma prevalence
- To describe the social and environmental factors associated with people with asthma
- To use publicly available data to suggest the priority areas of the intervention to control the asthma

## Data and Methods

### Data

We obtained model-based estimates of current asthma prevalence among adults aged 18 years or older for all 1314 census tracts in Cook County, IL. The asthma data, among many other health-related measures, are provided by the PLACES, Project of the Centers of Disease Control and Prevention (CDC 2021a) and stem from responses to the Behavioral Risk Factor Surveillance System survey (CDC 2021b). Specifically, asthma prevalence is quantified as the ratio between respondents who indicated having asthma and those who did not. Besides, we obtained census tract-level predictor variables from various resources: First, the CDC 2018 Social Vulnerability Index (CDC 2018), which quantifies social vulnerability to hazards like natural disasters or disease epidemics. The SVI is composed of four themes: 1) Socioeconomic Status, 2) Household Composition, 3) Race/Ethnicity/Language, and 4) Housing/Transportation. Second, we quantified spatial access to health care by calculating the density of medical centers using data provided by the 2015 Land Use Inventory of the Chicago Metropolitan Agency for Planning (CMAP, 2015). To avoid the modifiable areal unit problem (Openshaw & Taylor, 1979) which arises through aggregating the medical center locations to census tracts, we used the census tract-level average medical center density obtained by kernel density estimation (Silverman, 1986). We estimated density on a 30m grid, and employed a gaussian kernel with bandwidth chosen by Scott’s method (Scott, 1979). Third, we used daily census tract-level PM2.5 concentrations for 2016 (CDCPM25), which are modeled estimates using the United States Environmental Protection Agency’s Downscaler model (EPA). For each tract, we averaged the PM2.5 values across all 30m grid cells that intersect with the tract area, as well as across the entire timespan of hourly estimates. Lastly, we computed the census tract-level proportion of tree canopy area using the High-Resolution Land Cover, NE Illinois, and NW Indiana, 2010 dataset provided by CMAP. To conduct spatial analysis and mapping using geographic information systems (GIS), we obtained census tract polygon geometries as TIGER/Line Shapefiles from the United States Census Bureau. We joined all of our census tract-level variables to the geometries through their 11-digit FIPS codes.

### Spatial distribution of asthma

To illustrate the spatial distribution of asthma prevalence in Cook County, we utilize the purely spatial scan statistic (Kulldorff, 1997) with the normal probability model (Kulldorff, Huang & Konty, 2009), implemented in SaTScan™ software (Kulldorff, 2018). The spatial scan statistic identifies the most likely clusters of elevated asthma prevalence out of a set of candidate clusters. Each cluster *z* is a circle of radius *r*, centered on a candidate location, and multiple different radii are assessed per location. The set of candidate locations consists of the centroids of the *N*=1314 census tracts in Cook County, IL, therefore each centroid is the center of multiple candidate clusters of variable radii. Also, all census tracts whose centroid intersects with a circle are part of the respective cluster. The null hypothesis (*h*_*0*_) states that mean asthma prevalence inside the cluster is equal to outside, whereas the alternative hypothesis (*h*_*a*_) states that asthma prevalence inside the cluster is higher than outside (Kulldorff, Huang & Konty, 2009). We evaluate both, *h*_*0*_ and *h*_*a*_ using the likelihood ratio test in Equation (1):

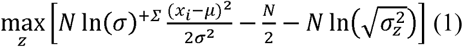

Where *x*_*i*_ is the asthma prevalence value at census tract *i*, μ the global mean, and *σ* ^*2*^ the variance.

Statistical significance of the most likely cluster(s) is evaluated using Monte Carlo simulation by randomly permuting the asthma prevalence values and their corresponding census tracts 999 times. For each simulation, we compute the log likelihood ratio, and the most likely cluster is noted. If the observed test statistic is within the top 5 percent among the simulated ones, we consider the cluster significant at the 0.05 alpha level. We restricted the clusters to contain a maximum of 50% of the underlying population to avoid overly large clusters that may be better represented as multiple smaller, disconnected clusters. We adopted the approach of Desjardins et al. (2020), Hohl et al. (2020), and Andresen et al. (2021), and mapped asthma prevalence for each census tract to illustrate variability within clusters identified by the spatial scan statistic.

### Spatial correlates of asthma

We determined significant predictors of asthma prevalence by employing ordinary least squares regression. First, we performed min-max normalization of all predictor variables to circumvent dependence on measurement units (Tan, Steinbach & Kumar, 2016). We avoided multicollinearity by computing the variable correlation matrix (Figure 1 in Appendix) and ensuring that variance inflation factors were below a recommended threshold of 2.5 (Cranley & Surles, 2002), indicating that collinearity among predictors did not lead to an inflation of variance. Therefore, our final regression model included the predictors SVI Theme 2, Theme 3, Theme 4, Tree Cover, Medical Center, and PM25. Our regression diagnostics included checking for heteroskedasticity by plotting residuals versus fitted values (Figure 2 in Appendix), as well as checking for normality by Q-Q plot (Figure 3 in Appendix).

**Figure 1.**
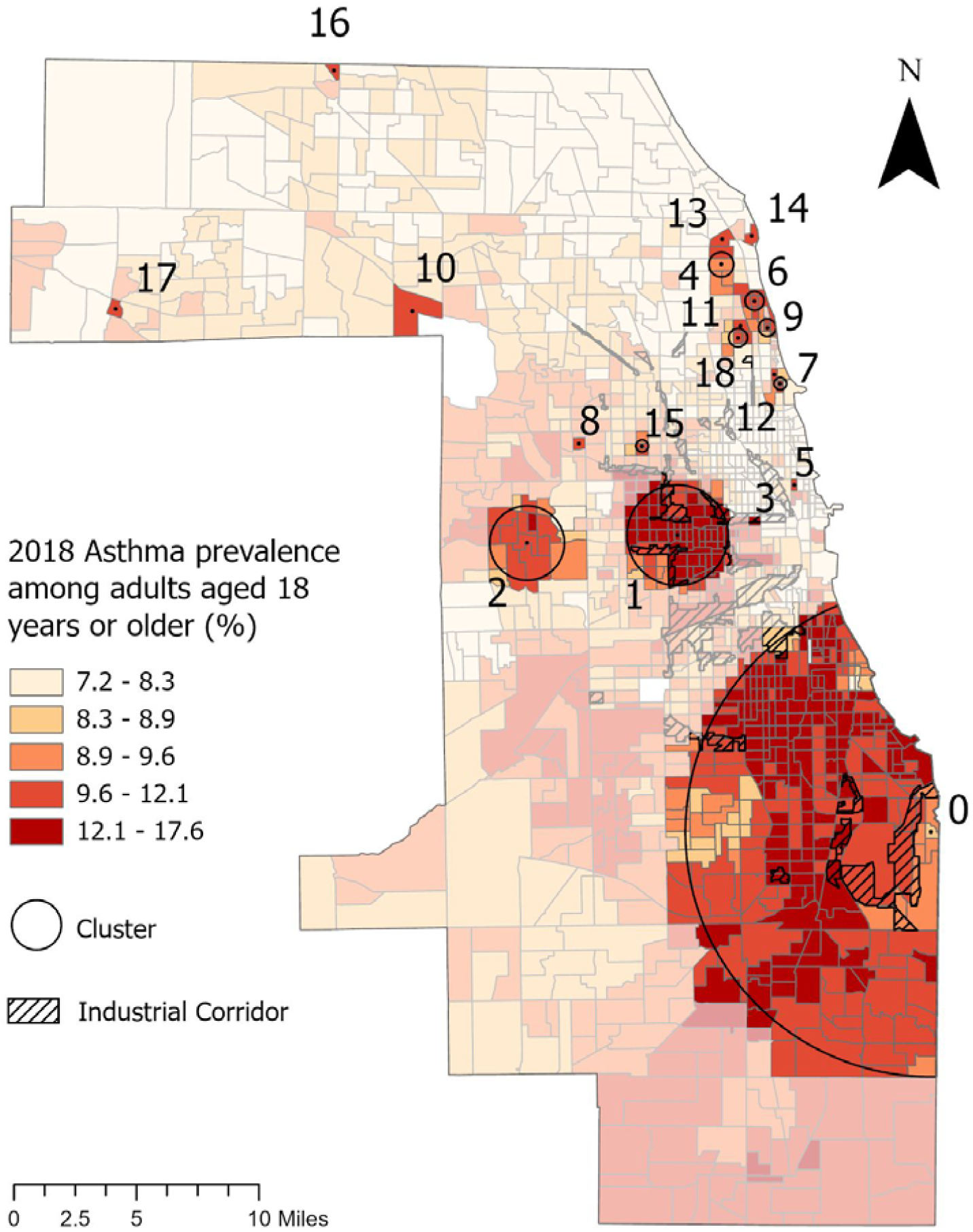
Clusters of elevated asthma prevalence.

**Figure 2.**
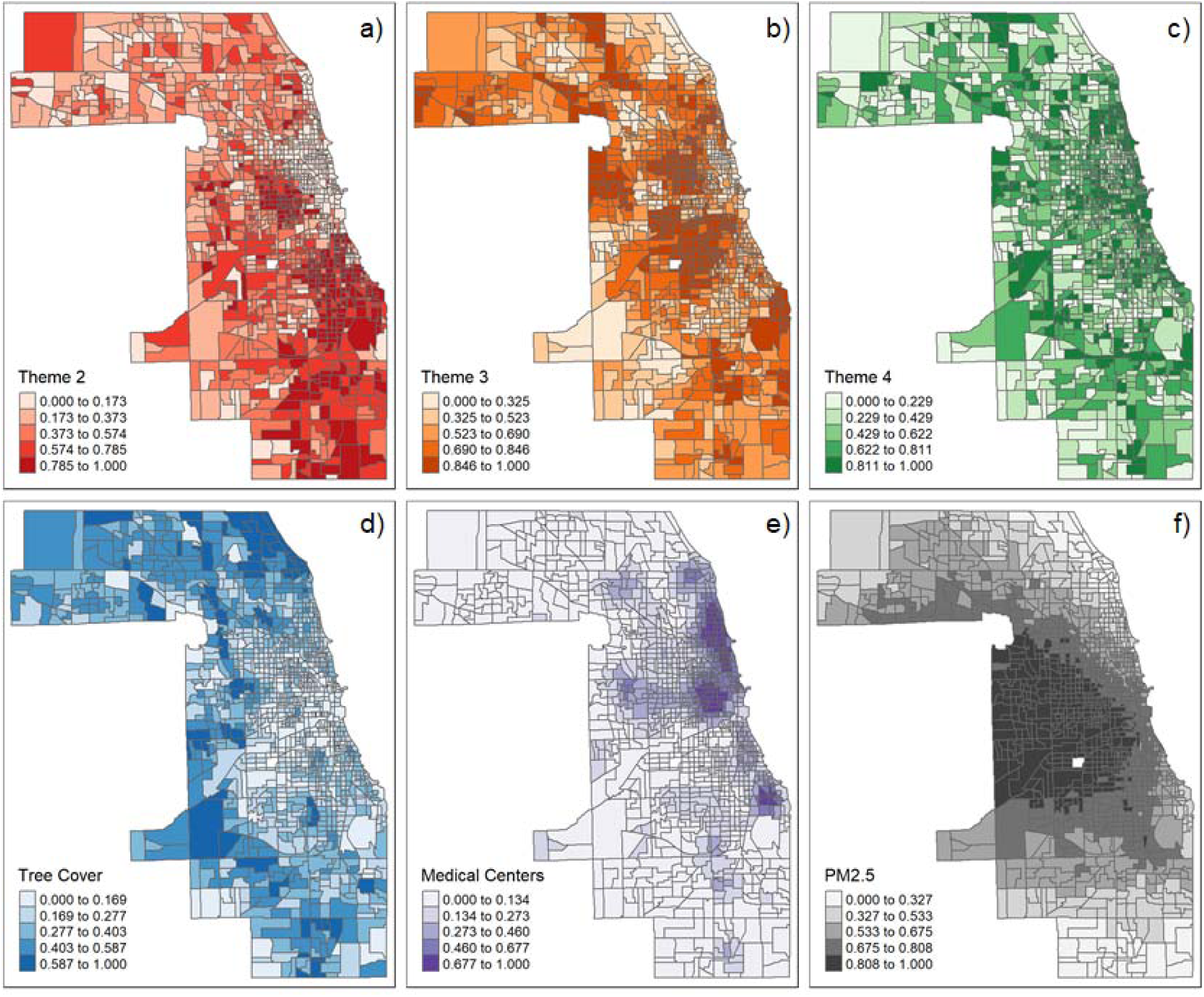
The spatial variations of Theme 2 (2a), Theme 3(2b), Theme 3(2c), Tree Cover (2d), Medical Centers (2e), and PM 2.5 (2F).

**Figure 3:**
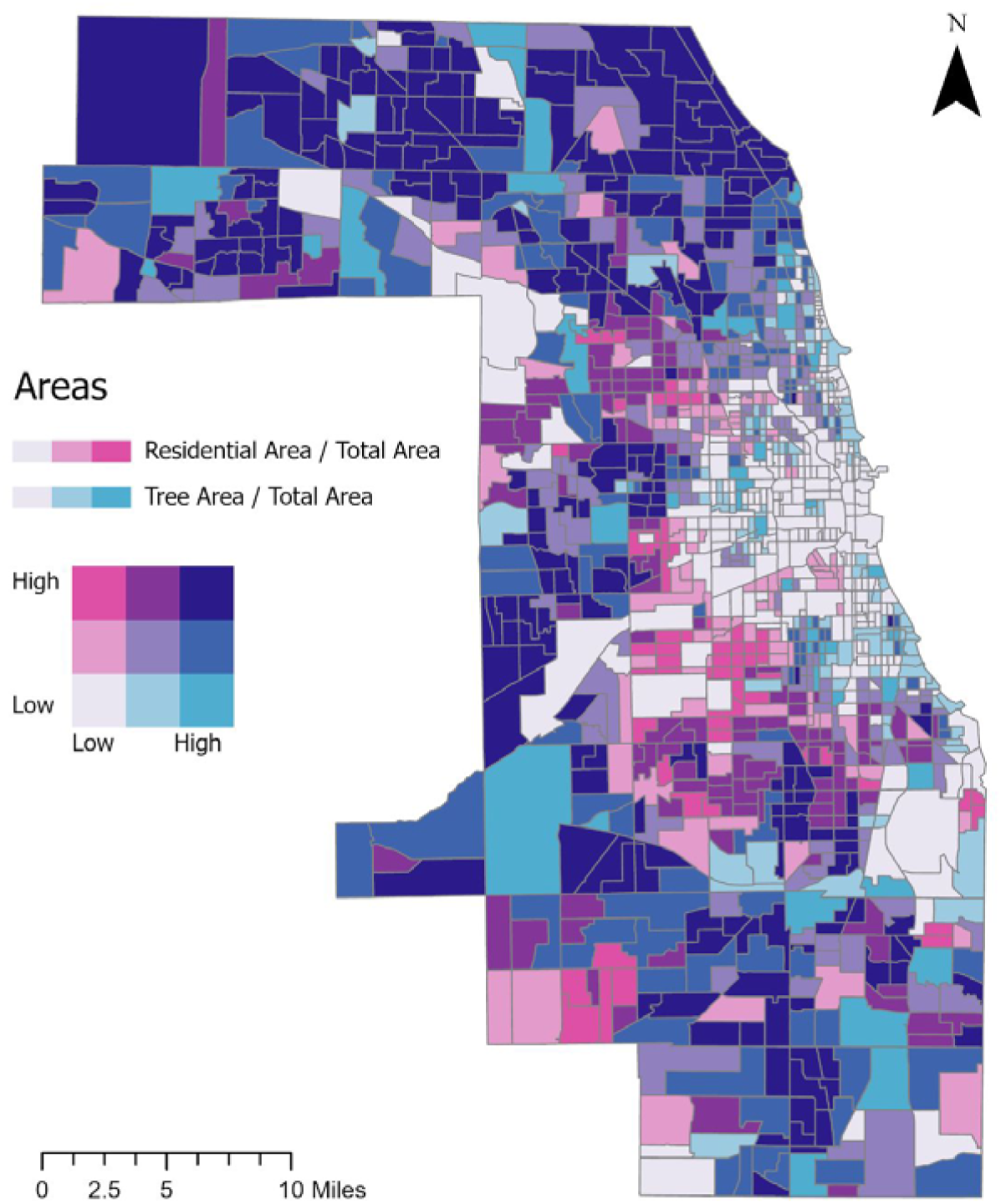
Bivariate Map of Residential Area Versus Tree Cover Area.

We further analyzed our OLS regression model to check for spatial autocorrelation of residuals, which constitutes a violation of the assumptions of OLS (Anselin, 2001). First, we visualized the spatial distribution of residuals by drawing them on a choropleth map of the census tracts in Cook County, IL (Figure 4 in Appendix). We tested for the presence of residual spatial autocorrelation by Moran’s I (Moran, 1950), and employed Lagrange Multiplier tests to determine whether autocorrelation is present in the dependent variable, or in the error term (Anselin, 2013). This sequence of diagnostics led us to account for spatial autocorrelation of OLS regression residuals by applying a spatial lag model with a queen’s spatial weight matrix.

All of our statistical computing was conducted using R (R Core Team 2017) and RStudio (RStudio Team, 2020) and we used ArcGIS Pro software for cartography (ESRI, 2020).

### Mapping tree coverage and residential area

To illustrate the ratio of residential land use to tree coverage per census tract, we draw a bivariate choropleth map of tree cover area and residential area at the census tract. The strength of the bivariate choropleth map lies in its concurrent display of two variables through the composite coloring of both variables (Lan et al., 2021). Bivariate maps have been utilized for visualizing the results of geographically weighted regression (Mennis, 2006), as well as illustrating smoking prevalence patterns (Liu, 2020). We use quantile classification for class breaks of both variables, since this is the preferred method for health-related data on choropleth maps (Brewer, 2006).

## Results

### Spatial distribution of asthma

The 2018 asthma prevalence among adults in Cook County, IL exhibits a fairly clustered spatial distribution (Figure 1). Using the spatial scan statistic, we identified 18 statistically significant clusters of elevated asthma prevalence, whereas the largest and strongest cluster (Cluster 0) is located in Chicago’s South Side. It has a mean asthma prevalence of 11.97% (Table 2), in proximity of the industrial corridor, an LLR of 1,186,958.95 and encompasses 297 census tracts. The second largest and strongest cluster (Cluster 1) is found west of downtown Chicago, with a mean prevalence of 13.26%, an LLR of 401,734.58, and it encompasses 61 census tracts. Cluster 2 is situated west of Cluster 1 near the village of Maywood and the planned manufacturing corridor. With an LLR of 20,580.59, it is substantially weaker than clusters 1 and 2, but has a mean asthma prevalence of 11.27. Several additional clusters are found in the northeastern part of the city along the shore of Lake Michigan. Table 1 shows important characteristics of the corresponding clusters in Figure 1. Table 1 is sorted according to the LLR, includes mean asthma prevalence inside the cluster (μ_in_), as well as outside (μ_out_), and the number of census tracts that are part of the cluster. Lastly, the table includes the p-value, which indicates cluster significance.

**Table 1.**
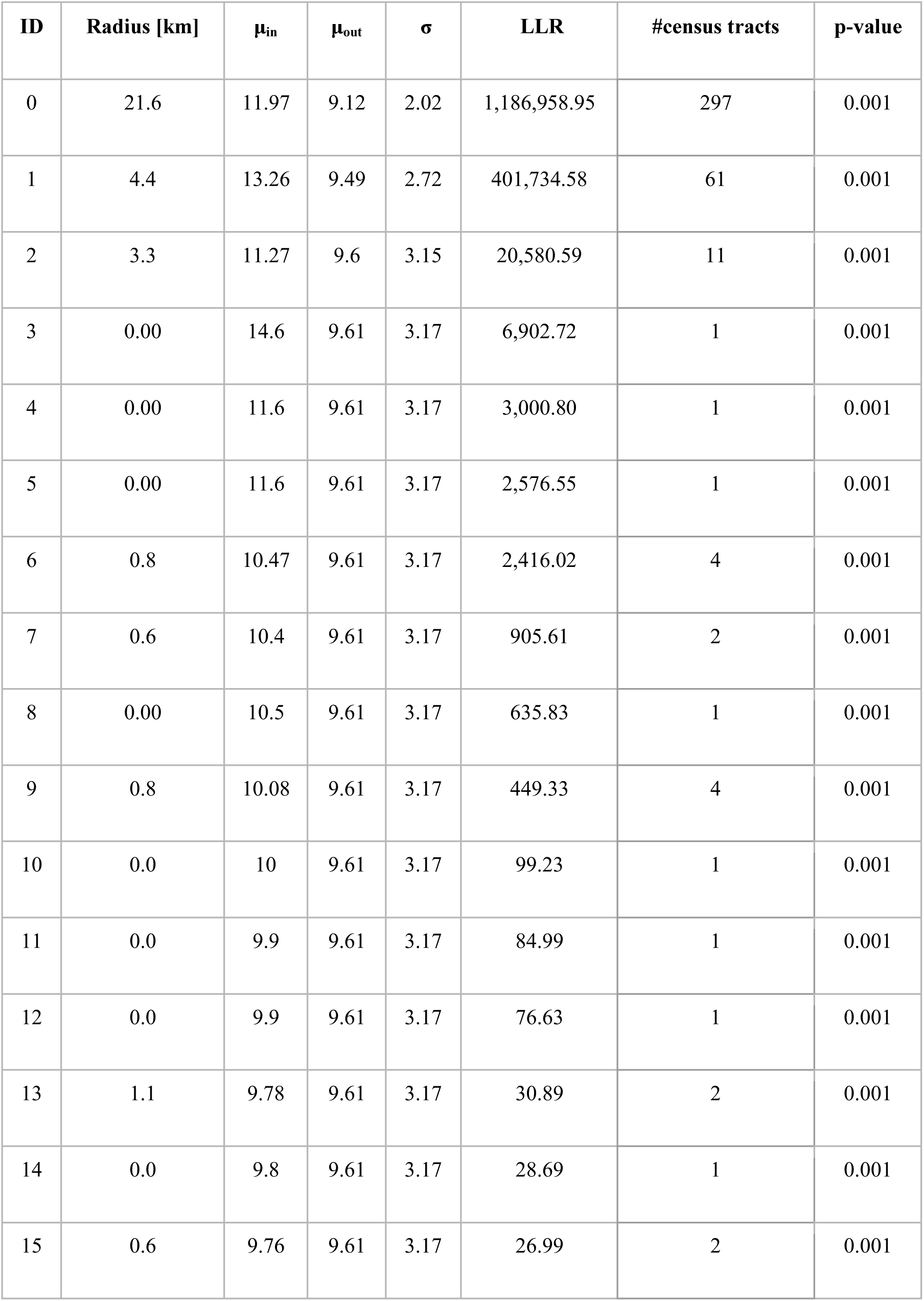
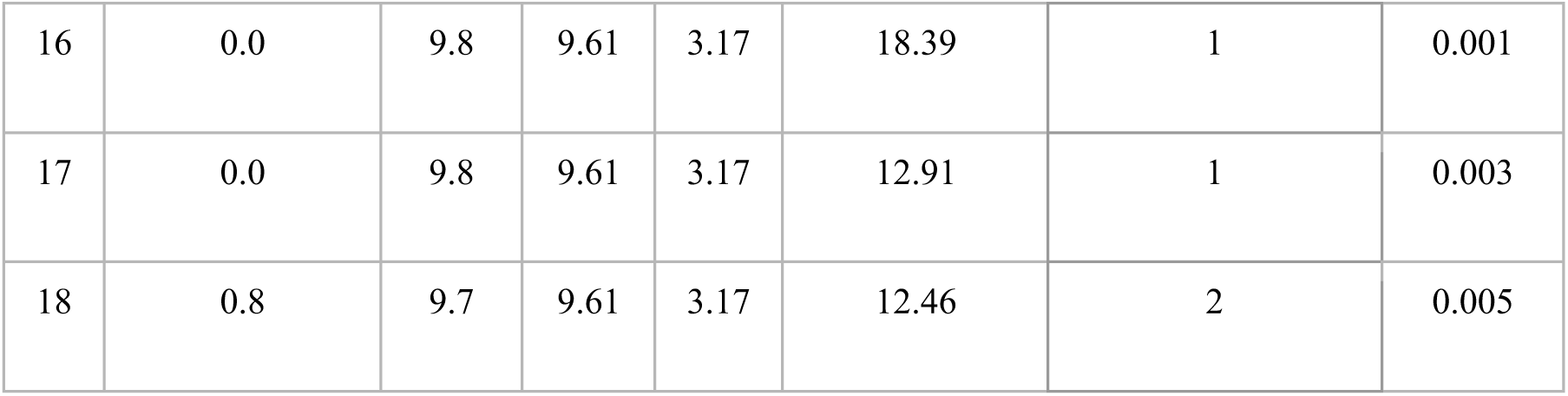
Clusters of elevated asthma prevalence. ID corresponds to cluster numbers in Figure 1.

**Table 2.** OLS and Spatial Lag Model results.

|  | OLS |  |  |  | Spatial Lag Model |  |  |  |
| --- | --- | --- | --- | --- | --- | --- | --- | --- |
|  | Estimate | Std. Error | t-value | p-value | Estimate | Std. Error | z-value | Pr(>\|z\|) |
| **Intercept** | 7.6 | 0.3 | 30.23 | 0.00 | 1.76 | 0.20 | 8.78 | < 2.2e-16 |
| **Theme 2** | 5.3 | 0.1 | 42.34 | 0.00 | 1.98 | 0.10 | 19.08 | < 2.2e-16 |
| **Theme 3** | -0.9 | 0.2 | -4.99 | 0.00 | -0.37 | 0.10 | -3.58 | 0.00035 |
| **Theme 4** | 0.7 | 0.1 | 5.01 | 0.00 | 0.59 | 0.09 | 6.71 | 1.97E-11 |
| **Tree Cover** | -1.7 | 0.3 | -6.4 | 0.00 | -0.58 | 0.17 | -3.50 | 0.000472 |
| **Medical Centers** | 0.7 | 0.2 | 4.09 | 0.00 | 0.25 | 0.11 | 2.28 | 0.022375 |
| **PM25** | 0.7 | 0.2 | 3.12 | 0.00 | -0.01 | 0.13 | -0.10 | 0.919097 |

### Spatial correlates of asthma

We found census tracts that exhibit high values of Theme 2 of the SVI conglomerate in the southern part of Cook County, as well as west of downtown (Figure 2a). These areas are therefore vulnerable in terms of household composition. Similarly, we found pockets of higher vulnerability in terms of Minority Status & Language (Theme 3) scattered throughout the county, with larger clusters southwest and west of downtown (Figure 2b). Theme 4 (Housing Type & Transportation) has a different pattern, where census tracts in the center of the city, as well as along Lake Michigan are more vulnerable (Figure 2c). However, the distribution of Theme 4 seems more random than the previous themes. Expectedly, areas with higher levels of tree cover are found at the fringe of the city (Figure 2d). Conversely, census tracts in the central parts of the city have lower levels of tree cover. Medical center density is very clustered, with areas of higher density found along the lake shore in the north, west of downtown, and southeast (Figure 2e). Lastly, the distribution of PM2.5 shows that areas in the western part of the city have lower air quality (Figure 2f).

The OLS regression revealed that household composition and disability (SVI Theme 2) wa positively associated with asthma prevalence (Table 2). Therefore, asthma prevalence is higher for census tracts that have higher proportions of people aged 65 or older, people 17 or younger, disabled people above 65, and single-parent households. In addition, we found a negative association of Minority Status and Language (SVI Theme 3), while housing type and transportation was positively related to asthma (SVI Theme 4). Therefore, census tracts that have higher proportions of multi-unit structures, mobile homes, higher levels of crowding, the asthma prevalence is higher.

The proportion of tree cover was negatively associated with asthma, meaning census tracts with higher tree coverage exhibited lower asthma. Then, the medical center density showed a positive association with asthma, meaning that asthma prevalence is higher where access to health care is high. Lastly, air quality (PM2.5) was positively associated with asthma. Overall model fit was good, with an R^2^ of 0.62.

The spatial analysis of the OLS residuals revealed that there is significant spatial autocorrelation in the model. Both, the choropleth map of residuals (Figure 4 in Appendix), as well as the Moran’s I test (I = 0.39, p = 0.00), confirm the presence of spatial autocorrelation of residuals. The Lagrange multiplier tests were highly significant for spatial lag, as well as for spatial error models (LMlag = 1054.4, p-value = 0.00; LMerr = 615.33, p-value = 0.00), whereas their robust form indicated that specification of a spatial lag model is appropriate (RLMlag = 459.19, p-value = 0.00; RLMerr = 20.174, p-value = 7.07e-06).

The spatial lag model mostly confirmed the results of the OLS model (Table 2). Findings include that living in census tracts that exhibit higher vulnerability in terms of household composition and disability (SVI Theme 2), as well as in terms of housing type and transportation (SVI Theme 4) is associated with higher asthma prevalence. Conversely, living in a census tract that exhibits higher vulnerability in terms of minority status and language (SVI Theme 3) is associated with lower asthma prevalence. In addition, areas with higher proportions of tree cover is associated with lower levels of asthma, whereas density of medical centers is associated with higher levels. Finally, air quality (PM2.5) does not have a significant association, as this variable became insignificant in the spatial lag model, which constitutes the only difference to the OLS model. As expected, the autoregressive lag coefficient indicated the presence of spatial dependence (ρ = 0.74, p = 0.00). Model fit of the spatial lag model (AIC = 3232.1) was higher than that of the linear model (AIC = 4359.5).

### Mapping tree coverage and residential area

Figure 3 illustrates the ratio of tree cover to residential land uses per census tract. While the extent of tree cover is high in low-density residential areas in southern Cook County, tree cover is low in the downtown and southwest part of downtown with the high-density residential areas. High dense residential areas with high-density tree cover are mostly in the North and Northwest of Cook County.

## Discussion

The asthma incidence varies across Cook county and it is higher in the South Deering residential neighborhood, the largest of the 77 community areas of the Chicago Metropolitan Area in South of Cook County. It is primarily a manufacturing industrial site, a small residential neighborhood existing in the northeast corner, and Lake Calumet taking up a large portion of the area. Also, the Roosevelt and Northwest industrial sites planned to maintain the corridors as an economic engine in the West of the County witness the second-highest asthma incidence. These findings can be used as evidence of geospatially varying respiratory health for urban planning decisions which aim to social and environmental interventions in reducing disparity in respiratory health and asthma prevalence.

Besides, the spatial association of asthma incidence with social and environmental factors was illustrated via using geospatial regression. The results show variation in asthma by age, disability, and family profile. The majority of people aged 65 and older, aged 17 and younger, with disabilities, and single-parent households located along South Lake Michigan and the West part of downtown are most at the risk of asthma. This calls for a cross-disciplinary study of land use planning in areas that are already identified with high asthma incidence. Meanwhile, the asthma incidence was elevated in overcrowding, multi-unit, and old buildings with poor air ventilation, and mobile homes along Lake Michigan though the asthma incidence was reduced among minorities and people with poor language proficiency in Northwest, Southwest, and West of Downtown. This might be because the communication barriers inhibit the minority with limited English Proficiency to communicate the asthma symptoms. Equal access to health care for people with limited language proficiency in equal need of health access asks for people centered health care that is timely, equitable, integrated and efficient. In the meantime, access to and use of asthma health services affect asthma management and control. Asthma incidence is higher where a large number of people with asthma are admitted to medical care. Quality of care is the degree to which health services for populations increase the likelihood of desired health outcomes. Besides the social determinants, environmental disparities have been associated with asthma incidence, encompassing spatial distribution of tree cover impacting air quality and reducing the impacts of exposure to air pollution as the large asthma incidence was found in districts with poor air quality in the West of Cook County and with a lack of tree cover throughout Chicago City. The geospatial regression illustrated the geospatial social-environmental determinants that might be associated with people with asthma and or people at the risk of asthma incidence.

Among those determinants, the neighborhood tree cover was related to better overall health by reducing the impacts of exposure to air pollution. The relationship between urban tree cover and residential land use in Cook County was studied. In Cook County, tree cover varies with housing density per census tract. While the densely housing areas left scarce plantable spaces in mostly Southwest and Northwest of downtown, canopies are mainly isolated and spatially segregated from the built-up framework in the low-density residential districts in South, Southwest, Northwest of Cook county. The practices of urban planning and landscaping in new development and renewal areas should take into account some measures to augment tree cover, especially in districts with the largest asthma cluster.

Specific to asthma incidence, the result of this study aligns with factors used in the previous study (e.g. Lan et al., 2018; Edwards et al., 2006) that land uses, lack of green space, impacts of exposure to air pollution, and residency near heavy industry were associated with respiratory diseases.

There are five main limitations in this study. First, the asthma variable is the ratio between respondents who indicated having asthma and those who did not. The diagnosis of asthma is self-reported and is not physician-confirmed. Secondly, this study does not include the impacts of micro-community environments and indoor residential environments. Future research can capture these impacts with variables to represent spatial features of these two levels. Third, we employed the spatial scan statistic, which produces clusters of circular shapes. However, the circular cluster assumption may not hold true in a spatially heterogeneous area. The spatial scan statistic has been extended to address this issue (Kulldorff et al., 2006; Tango & Takahashi, 2005; Duczmal et al., 2007), but using circular clusters as implemented in the SaTScan™ software is still a standard practice in spatial analysis. Fourth, the retrospective study design limits our capacity to establish causal relationships. However, our analysis can guide future research efforts to identify neighborhoods with geographic disparities and the associated underlying causes of elevated asthma prevalence. Fifth, our data is aggregated to census tracts, which may cause the modifiable areal unit problem (MAUP; Openshaw and Taylor 1979), resulting in complications with our statistical analyses.

## Conclusion

The main finding of this study is that the spatial distribution of asthma incidence overall varies based on demographic profile, green space, transportation, land use planning, and language barrier. Accordingly, suggestions for planning intervention include 1. Balance vegetation coverage and residential density 2. Protecting and equal distribution of vegetation cover to remove the air pollution especially for improving air quality in the residential areas in the proximity of the industrial corridors 3. Finding solutions for old multi-use, and crowded buildings in high-density urban areas with poor air quality.

This cross-sectional study attempts to explore the socio-environmental variables associated with asthma prevalence while it is limited to the public data. Further study is needed to discover the causality between the built environment and asthma prevalence. More specific areas such as manufacturing and high-density urban areas can be thoroughly explored in further study. Longitudinal rather than cross-sectional studies can be used to unravel the causal relationship between built environmental factors and respiratory health outcomes.

## Data Availability

Data used in this study are publicly available data.

